# Sequence Analysis as an approach to characterize variables that unfold over time: implementation and practical considerations for epidemiologists

**DOI:** 10.1101/2024.06.18.24308957

**Authors:** Lucia Pacca, Kristina V Dang, Leah R Koenig, Catherine dP Duarte, S Amina Gaye, Amal Harrati, Anusha M Vable

## Abstract

Characterizing longitudinal trajectories of social exposures or health outcomes is a persistent challenge, but can be accomplished with sequence analysis, a data-driven approach that can differentiate timing, order and duration of events. We present practical guidance on implementing sequence analysis for epidemiologists with the goal of providing clear advice on decision points and tradeoffs.

We Introduce the three main steps of sequence analysis: (1) coding longitudinal processes as trajectories of ordered events for a set of individuals, (2) measuring dissimilarity between individual trajectories, and (3) performing cluster analysis to group similar trajectories. Each of these steps presents researchers with several decision points, such as data cleaning rules, options for evaluating sequence dissimilarity, and choices of clustering algorithms to group trajectories. After outlining each of the sequence analysis steps, we provide an applied example of sequence analysis in which we create and group transition-to-retirement trajectories from age 51-75 for a sample of 9,189 Health and Retirement Study participants using self-reported employment information, then estimate the association between transition-to-retirement groups and self-rated health.

Our paper seeks to guide epidemiologists through the analytic decisions and implementation challenges of sequence analysis as this approach is increasingly implemented and undergoes methodological advances.

## INTRODUCTION

Lifecourse epidemiology, an established subfield of epidemiology, has been important in advancing a more nuanced examination of the complex relationship between socio-economic position (e.g., education, income, work) and health over time^1,2^. To-date, a lifecourse focus has been particularly useful for documenting early-life physical and social determinants of chronic diseases with long latency periods, such as hypertension and diabetes^3–5^. Research within the subfield is increasingly moving beyond exploration of single point-in-time exposures during critical or sensitive periods in early-life to considering the cumulative health effects of exposures^2,6^.

Socio-economic factors evolve and interact in complex ways across the life course^7^. The timing, duration, and order of these factors varies considerably across individuals in ways that may differentially impact health outcomes. This heterogeneity cannot be adequately captured with approaches that restrict exposure operationalization to specific windows of time or average over multiple time points. Approaches that describe, as opposed to obscure, this heterogeneity are necessary.

Sequence analysis is one such approach facilitating the characterization of lifecourse trajectories^8^. As a data reduction technique, sequence analysis simultaneously considers the timing, duration, and order of longitudinal exposures, revealing common patterns in the data which can be used to classify individuals into distinct groups^9^. Originally introduced in biology to analyze sequences of proteins and DNA, sequence analysis was first integrated into the social sciences by Abbott and Forest in 1986^10,11^. Since then, there have been many applications of sequence analysis in social science research, with growing interest in social epidemiology and healthcare^12–16^.

A typical application of sequence analysis involves three steps: (1) creating individual trajectories of ordered events, (2) quantifying trajectory dissimilarity, and (3) performing cluster analysis to group similar trajectories. Implementing sequence analysis, however, can be challenging since each of these steps presents researchers with several analytic choices. For example, Step 1 of creating individual trajectories of ordered events can involve intensive data cleaning decisions. Step 2 of quantifying trajectory similarity requires informed selection from among a variety of existing algorithms, the comparative strengths of which can be unclear. In Step 3, there are also several options for implementing cluster analysis and selecting the final number of trajectory groups. That there are several subjective decision points along the way can be daunting, particularly for those applying sequence analysis for the first time.

To support epidemiologists in navigating these challenges, we present a systematic guide on sequence analysis implementation. Using the publicly available Health and Retirement Study data, we provide an applied example wherein we use sequence analysis to identify transition-to-retirement trajectories then estimate their association with later-life health. We conclude by orienting readers to the available packages and options in two types of statistical software (R and Stata). Accompanying code is available on the following GitHub page [redacted for peer review].

## OVERVIEW OF THE STEPS OF SEQUENCE ANALYSIS

### Step 1: Creating individual trajectories as sequences of ordered events

The idea behind sequence analysis as applied to lifecourse data is to represent life course processes as “sequences’’ (i.e., individual-level trajectories) of ordered “states” (i.e., mutually exclusive events) over a defined observation period. That is, for each study participant in a given sample, each time point (e.g., year, month, week, day, hour) within the observation period is assigned to a mutually exclusive categorical state. As a simplified example, say a researcher is interested in empirically characterizing employment sequences. For each study participant in the sample (Figure 1, rows), they would assign a mutually exclusive employment state (employed, unemployed, retired) to each time point (per 1-month) over the chosen observation period (10 months; Figure 1, x-axis).

**Figure 1.**
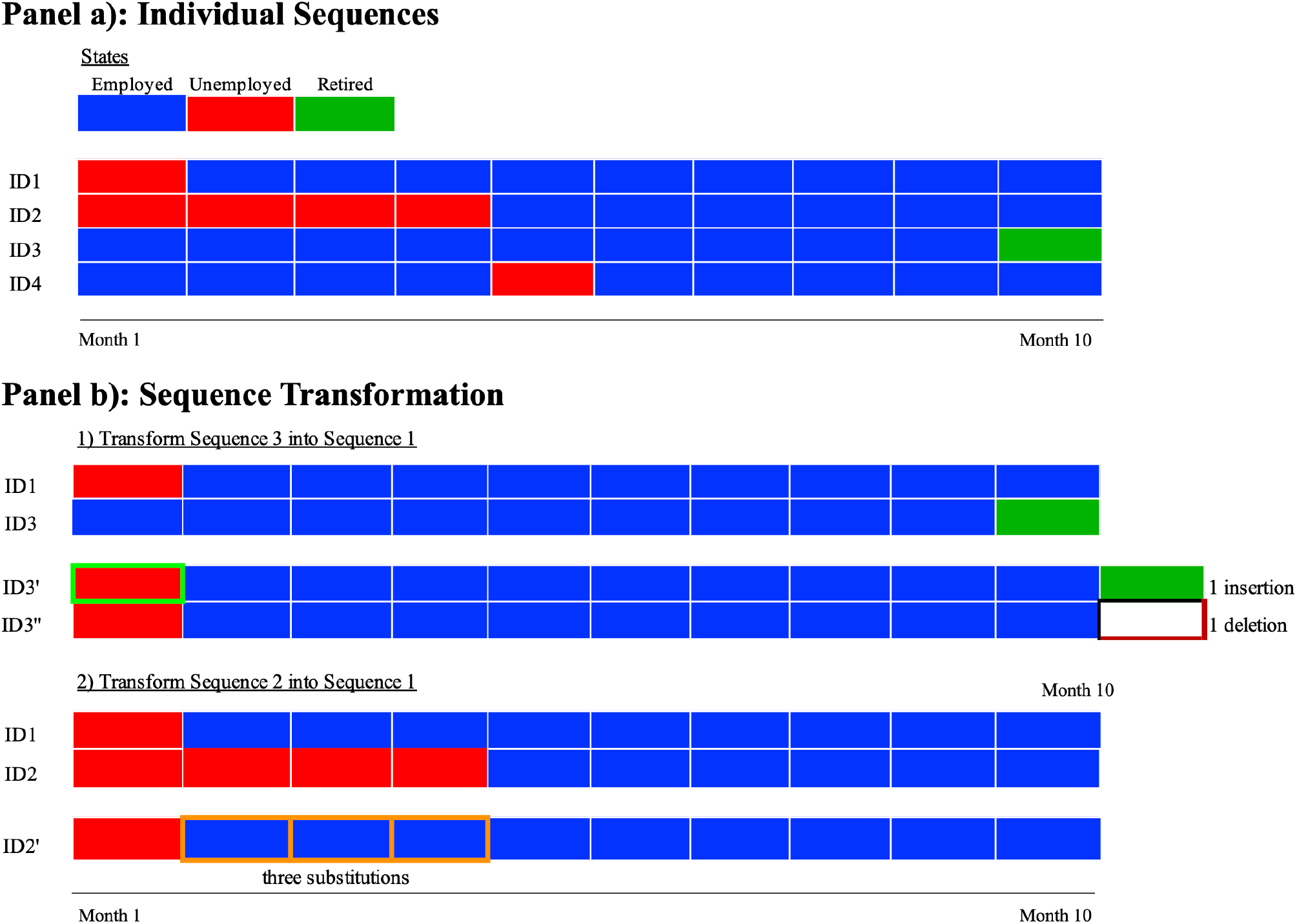
– Individual sequences and their transformations. This figure presents a simplified example of four individual employment trajectories (Panel a) and of the basic edit operations used in sequence analysis to compare trajectories (Panel b). Employment trajectories are sequences of ordered, mutually exclusive categorical states (in our example, we chose three states: employed, unemployed and retired) over a 10 months period. The four listed sequences differs based on the experienced states as well as their timing, duration and order. Sequence analysis measures sequence dissimilarity by assigning “costs” corresponding to edit operations (insertions, deletions or substitutions) to turn each sequence into all the other sequences in the data. In Panel b, transformation 1) transforms ID1 into ID3 by first inserting “unemployed” state in month 1, and then deleting “retired” state in month 11 (the insertion extended ID3 by one month). Transformation 2) transforms ID2 into ID1 by substituting “unemployed” state with “employed” state in months 2, 3 and 4. While insertion and deletion (indel) operations shift sequences and distort timing of events, substitutions preserve timing.

When analyzing large datasets, this process usually results in a multitude of unique sequences, which may differ from each other not just by experienced states, but by the order, duration and timing of states as well^17^. These four aspects, which have been identified as critical for the sociological meaning of sequences, are defined as follows:

1. Experienced states - the mutually exclusive events, experienced at different time points in the sequence
2. Order - the sequential arrangement of states over the total observation period
3. Duration - the number of consecutive time points spent in each state over the total observation period
4. Timing - the particular time point at which each state onsets over the total observation period

For example, in Figure 1 (Panel a) ID1 and ID2 experienced the same states (employed and unemployed) over the 10-month time period with similar timing (start unemployed state at month 1) and order (unemployed to employed), but different duration (1 month vs. 4 months of unemployed). ID3 experienced different states (employment and retirement) than ID1 with different timing, but similar duration (9 months of employment). Finally, ID4 experienced the same states (employed and unemployed) as ID1 with similar duration of unemployment (1 month), but different order (ID1: unemployment, then employment vs. ID4: employment, unemployment and then employment again).

Epidemiologic research often involves the use of population-level datasets comprised of thousands of individuals followed over hundreds of time points. Creating individual sequences for such a large number of participants and long time period is a time-consuming process, as real-world longitudinal datasets usually include a proportion of missing information and can have inconsistencies^18^. Generating the trajectories requires defining a set of data cleaning rules, which will be illustrated more in detail in the applied example.

### Step 2: Quantifying trajectory dissimilarity across participants

Sequence analysis allows comparing thousands of observed sequences by producing a dissimilarity (or distance) measure for all pairs of sequences in a given dataset. There are several options for algorithms to quantify pairwise sequence dissimilarity, most of which involve some measure of the effort necessary to transform one sequence into another.

#### Transformation Operations

Optimal Matching (OM) is the most used method to compare sequences^10,11^. OM calculates dissimilarity as the minimum effort necessary to transform one sequence into another through three kinds of transformation operations, each of which is assigned a “cost”:

1. Insertion: inserting a state into a sequence
2. Deletion: deleting a state from a sequence
3. Substitution: replacing one state with another state

Figure 1 (Panel b) illustrates these transformations, using the sequences first introduced in Figure 1. Transformation 1) illustrates the insertion and deletion (“indel”) operations. Specifically, ID3 is transformed into ID1 by applying one insertion (employment in month 1, ID3’) and one deletion (retirement in month 11, ID3’’). Transformation 2) illustrates the substitution transformation. Specifically, ID2 is transformed into ID1 by applying three substitutions (replacing unemployment with employment in months 2-4, ID2’).

#### Assigning Costs

There are several options for selecting costs. For substitution cost selection, the three most common options are: 1) constant substitution costs (e.g. all substitutions have a cost of 2); 2) variable substitution costs chosen by the researcher, typically motivated by a hierarchy between states; and 3) variable data-driven substitution costs based on transition rates between states such that more frequent transitions have lower costs. Costs are represented by a substitution cost matrix, whose rows and columns correspond to the number of states.

Indel costs are most commonly set equal to ½ the maximum substitution cost but can also be customized if desired. Because insertion and deletion transformations cause a time distortion through shifting sequences (Figure 1, Panel b), the recommendation is either not to use them, or to set them higher (e.g., ⅔ the maximum substitution cost) when the timing of events is the main dimension of interest^17,19^.

#### Obtaining Total Costs

Once costs have been assigned, the OM algorithm selects the minimum total cost (combination of indel and substitution operations) necessary to transform each sequence into all the other sequences in the data. Applying this algorithm produces a distance (or dissimilarity) matrix, whose number of rows and columns corresponds to the number of individual sequences. Within this square, symmetric matrix, higher values indicate more dissimilar sequences. Besides Optimal Matching, many other techniques to calculate sequence dissimilarity have been developed and are now available^17,20^. Many of these options are variations of the traditional OM^21–23^. Because the sensitivity of each of these variations to the different sequence characteristics (order, duration, and timing of events) changes, the choice of which measure to use should be based on the specific research question. Table 1 summarizes the main dissimilarity measures recommendations from Studer et al. (2016)^17^ based on their sensitivity to order, duration, and timing of events. For example, if capturing timing is the main interest, the top recommendation is to use the Hamming distance^24^, which calculates sequence dissimilarity as the number of total substitutions and does not allow for indel costs (Table 1). However, while OM can be used with sequences of different length, Hamming requires all sequences to be the same length.

**Table 1.** – Recommended dissimilarity measures based on sequence characteristics.

| Characteristic of interest | Dissimilarity measure | Description | Sequences of different length |
| --- | --- | --- | --- |
| Duration | Optimal Matching (OM) | Traditional Optimal Matching; measures the dissimilarity between sequences as the minimum cost combining substitution and indel operations | Yes |
|  | Optimal Matching of spells (OMspell) with high "expansion cost" | Variation of Optimal Matching calculating substitution costs between sequences of spells (consecutive time periods spent in one state); higher expansion costs make the measure more sensitive to duration | No |
| Timing | Hamming (HAM) | Measures the dissimilarity between sequences as number of substitutions between non-matching states; does not use *indel* costs and requires sequences to be the same length | Yes |
|  | Optimal Matching (OM) with high *indel* costs | Traditional Optimal Matching; increasing indel costs makes their use more rare, resulting into a lower distortion of time | No |
| Order | Optimal Matching of transitions (OMtran) | Variation of Optimal Matching where substitution costs are based on transitions between states rather than on states themselves | No |
|  | Optimal Matching of spells (OMspell) with low "expansion cost" | Variation of Optimal Matching calculating substitution costs between sequences of spells; lower expansion costs make the measure less sensitive to sequencing | Yes |

This table shows the main recommended dissimilarity measures for sequence analysis based on the trajectory characteristic of interest (duration, timing or order of events) based on the comparative review by Studer (2016). For some of the listed dissimilarity measures, customizable options are available, which can change their sensitivity to timing, order and duration of states. For example, for traditional Optimal Matching, higher indel costs increase its sensitivity to timing of states. Similarly, OMspell gives the option of setting an “expansion cost” (cost of expanding one spell by a unit). Higher expansion costs make the measure more sensitive to duration compared to order of states.

**Table 2.** – Cluster quality measures commonly used in sequence analysis

| Measure | Description | Range | Min/Max |
| --- | --- | --- | --- |
| Average Silhouette Width (ASW) | Based on the similarity of the assignment of a trajectory to a given cluster, the ASW compares the average weighted distance of an observation from the other members of its group and its average weighted distance from the closest group. | $[-1, 1]$ | Max |
| Calinski-Harabasz index (CH) | Looks at the sum of squared distances within the partitions, and compares it to that in the unpartitioned data, taking account of the number of clusters and number of cases. | $[0; +\infty[$ | Max |
| Duda-Hart cluster-stopping rule (DH) | Similar to the Calinski-Harabasz index, but specific for hierarchical clustering. Comparing the sum of squares in the next pair of clusters to be combined, before and after combining. Displays two indices, $Je(2)/Je(1)$ and its pseudo-T squared. | $[0; +\infty[$ | $Je(2)/Je(1)$: Max;<br>pseudo-T squared: Min |
| Hubert's & Levin C index (HC) | Compares the partition obtained with the best partition that could be obtained with this number of groups and this distance matrix. | $[0, 1]$ | Min |

This Table presents four cluster quality measures commonly used in sequence analysis to choose the “optimal” number of trajectory clusters. A brief description of the measures and the range of their possible values are displayed in columns 2 and 3. Column 4 indicates whether each measure should be minimized (Min) or maximized (Max) to identify the best quality cluster solution. AWS and CH are available in Stata and R. DH is only available in Stata, while HC is only available in R.

### Step 3: Performing cluster analysis to group similar participant trajectories

Based on the dissimilarity matrix, cluster analysis is typically utilized to identify groups of similar sequences. Several clustering algorithms can be used to conduct cluster analysis. The two clustering methods most used in sequence analysis are Ward’s hierarchical agglomerative clustering^25^ and Partitioning Around Medoids (PAM)^26^.

#### Hierarchical Agglomerative Clustering

Hierarchical agglomerative clustering starts by considering each individual trajectory as its own cluster. At each iteration, the two least dissimilar groups (or initially trajectories) are grouped together, until all the observations form a single cluster. This creates a hierarchy of clusters, which can be represented as a tree. One can then choose the final number of clusters by “cutting” the tree in correspondence with a specific number of partitions. An advantage of hierarchical, agglomerative clustering is its tendency to produce clusters of similar size. However, a potential limitation of this method is its lack of flexibility, since the clusters that are created at each step can be combined with other clusters, but never split^8^.

#### Partitioning around Medoids (PAM)

A more flexible approach is the PAM algorithm, an extension of the k-means algorithm. This method finds representative sequences within the data set, called medoids, and creates the clusters through the association of each sequence to its closest medoid based on the distance matrix. The goal is to minimize the sum of the dissimilarities of the observations to their closest representative sequence.

#### Alternative Clustering Methods

Though less common, other clustering methods have been used to group trajectories in particular circumstances. Divisive clustering, a recently introduced and promising approach, allows specifying clearly the rules for cluster assignment^27,28^. Fuzzy clustering, which allows cases to belong to more than one cluster in varying degrees, is a promising and more flexible approach compared to “crisp” clustering, since it allows classifying sequences as hybrids (i.e., mixture between more than one cluster)^29^.

#### Selecting the optimal number of clusters

For any of these methods, the choice of the final number of clusters is made by the researcher. Several cluster quality measures can be used to assist in making this choice, with high-quality generally indicated by the number of clusters that maximize within-cluster homogeneity and between-cluster heterogeneity^30^. A description of the most used cluster quality measures is presented in Table 1. Although these measures can provide useful guidance to select the desired number of clusters, they can be integrated with other criteria, such as minimum cluster size, interpretability, and association of clusters with other variables that are expected to be related to the typologies of trajectories^31^.

## IMPLEMENTATION AND APPLIED EXAMPLE (TRANSITION TO RETIREMENT)

In this section, we provide guidance on implementation of the three main steps of sequence analysis through the identification of transition-to retirement trajectories in the Health and Retirement Study, a publicly available dataset of older U.S. Americans. We then use the trajectory clusters as exposures in a regression analysis aimed at investigating the association between transition-to-retirement trajectories and self-rated health. The R and Stata code for this analysis is available at the following GitHub page [redacted for peer review].

### Step 1: Creating, cleaning and visualizing individual sequences

To begin characterizing the transition-to-retirement trajectories, we first defined our total observation period as the 15 years from when participants were aged 51 to 75, which has been shown to capture the relevant range of ages over which most US residents tend to transition to retirement^32^. Although the data were collected biennially in calendar years, we switched the time scale to age and used retrospective information, when available, to determine employment status during non-survey years. Guided by our research question, any restrictions posed by the available data, and practical considerations regarding level of detail for the didactic purposes of this paper, we then chose the following 5 mutually exclusive employment states: (1) employed full-time, (2) employed part-time, (3) retired, (4) disabled, or (5) out of work. Too few states might not capture the heterogeneity of lifecourse trajectories, while too many states (e.g, more than 10 states) could result in clusters that are too heterogeneous to be interpretable. We created individual trajectories for our sample (N=9,189 participants from HRS entry cohorts 1992 - 1998) by assigning each year to one of the five mutually exclusive states.

#### Missingness decisions

When creating individual-level trajectories, an important consideration is the presence of missing data. Three main types of missingness are found in longitudinal sequence data: [1] initial missing data, due to individuals entering the sample at different time points; [2] terminal missingness, due to loss to follow-up; and [3] intermediate missing gaps, due to data not being reported or collected for some of the time periods. There are several options to treat missing time points. A first option is to exclude participants with a certain percentage of missing time points, though this may induce selection bias^33,34^. A second option, only applicable to terminal missingness, is to retain sequences of different length. A third option is to code missingness as an additional state, which can be applied to all three types of missing data and is particularly useful when missingness may be informative. Finally, a fourth option is to impute missing gaps. A chained multiple imputation procedure specifically developed for longitudinal data can be used, which fills missing gaps by using prior and subsequent state as key predictors, plus additional predictors^35^.

In our applied example, we decided to create an additional state, labeled “unreported”, to characterize internal missing gaps, since unreported employment status may be informative (e.g., indicative of periods of unemployment or more unstable work). Initial missingness was also coded as a separate state (“left missing”), indicating individuals entering the survey older than 51 and not reporting their retrospective employment information. Because we excluded respondents who were lost to follow up, there was no terminal missingness. The combination of five chosen states and two missing categories produced a total of seven final states.

#### Additional Data Cleaning Decisions

In addition to decisions related to missingness, there are usually other data cleaning decisions made when creating longitudinal sequences. For example, we can decide to collapse states that are rare and may be included in wider categories. In our example, the “out of work” category incorporates three states that are infrequently presented in the dataset: homemaker, unemployed, and temporary leave (2.87% of total states). Moreover, because participants reported information on previous employment every two years, we found some inconsistencies between information reported in different years. For inconsistent data between surveys, we defined states based on the most frequently reported information (mode) between surveys or, when more than one mode was present, on the most recently reported information. Because creating individual sequences may require defining several cleaning rules, we recommend listing these rules as part of a paper’s supplemental material for transparency and reproducibility.

#### Visualization using index plots

Once defined, all the individual-level trajectories can be simultaneously visualized using a sequence index plot, where each row along the y-axis corresponds to a unique participant and each column on the x-axis to each time point over the total observation period (in this case years from age 51 - 75), with each color representing a different state (Figure 4). Figure 2 displays the sequence index plot for the 9,189 participants included in our example, among which there were 4,649 unique trajectories.

**Figure 2.**
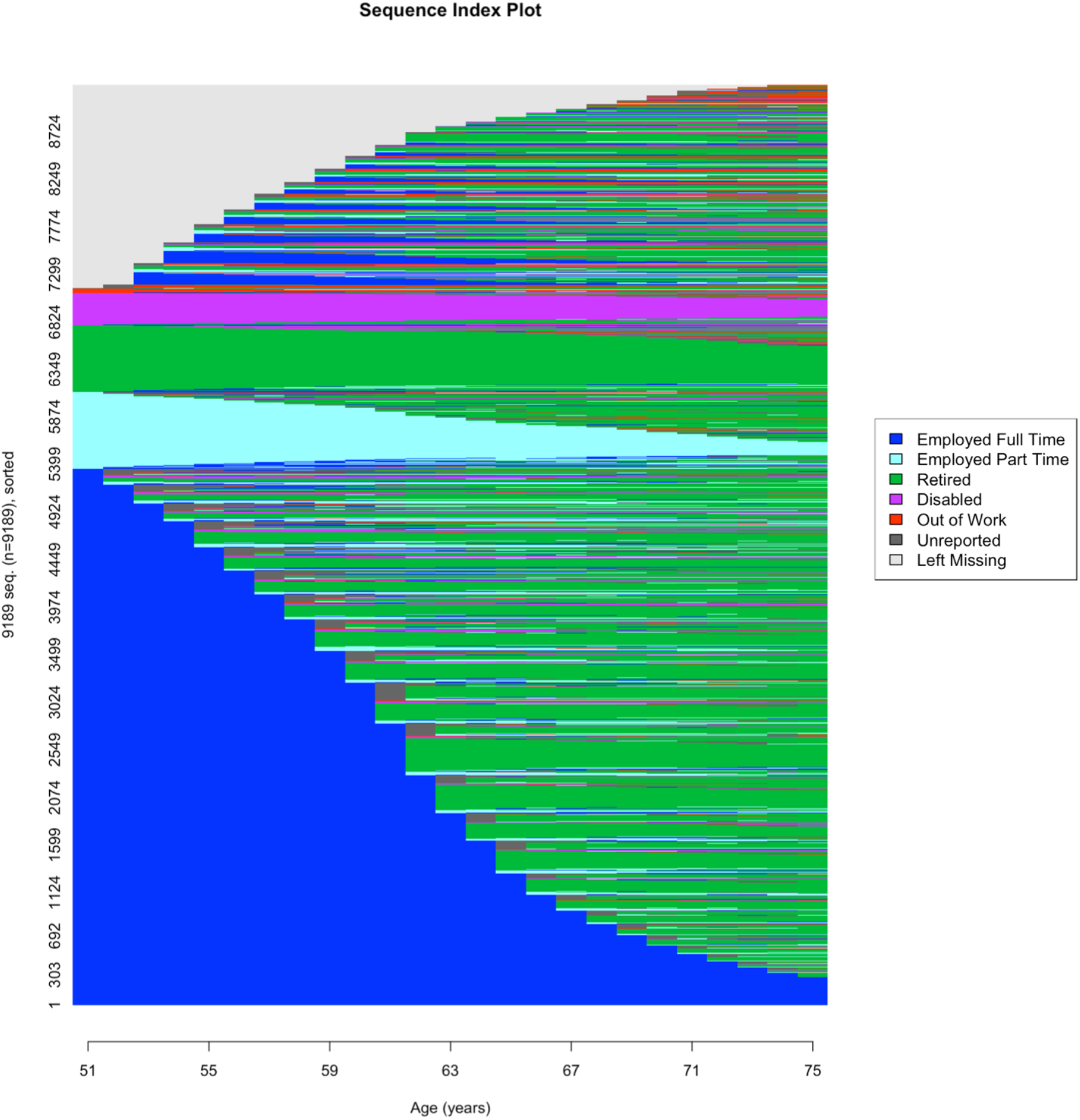
– Sequence Index Plot visualizing transition to retirement trajectories. This figure represents all the transition-to-retirement trajectories for the Health and Retirement Study participants eligible for our didactic example using a sequence index plot. The sequence index plot is the most complete graphical representation of sequences and can be helpful to graphically visualize heterogeneities. It represents all the individual trajectories (N=9,189, y-axis) over the chosen period (age 51-75, x-axis). The graphic shows that there are some repeated sequences in our dataset. For example, on the bottom of the plot, we can observe a share of participants (represented by dark blue lines) who were consistently employed full-time during the observation period. Most of the participants, however, experienced at least one transition between employment states. In our dataset of 9,189 participants, there were 4,649 unique trajectories.

### Step 2: Sequence comparison

First, we set substitution and indel costs based on transition rates. Although some states may be qualitatively more similar to each other, we could not establish a hierarchy that would guide our arbitrary cost assignment. We therefore exploited the transitions between states in the data, setting substitutions of states with high transition rates as less costly. This produced a symmetric substitution cost matrix (Table S2), with costs ranging between 2 and 0.

Next, we selected our preferred dissimilarity measure. Because we were interested in timing (e.g., early vs. later retirement) more than duration or order of events, we selected the Hamming distance. Since we categorized missingness as either “unreported” or “left missing” state, all trajectories were the same length; therefore, we did not need to use insertion and deletion operations to compare sequences. Applying the Hamming distance produced a dissimilarity matrix comparing each trajectory to all the other trajectories in the data (Table S4).

### Step 3: Clustering

To select between a hierarchical clustering or PAM approach, we compared a selection of cluster quality measures for the “best” partitions identified by the two different algorithms (reported in the manuscript’s supplement, Table S3). We chose to employ PAM as it displayed, overall, better cluster quality.

To identify the final number of clusters, we then graphically visualized the Average Silhouette Width (ASW) and Hubert’s C (HC) measures for a set range of cluster solutions (from 2 to 20 clusters, Figure 3). According to both indicators, 7 clusters represented the highest quality solution. Before choosing 7 clusters as the final solution, we graphically visualized them for qualitative evaluation (Figure 4) and looked at clusters’ size as well. The number of participants in each of the 7 clusters ranged between 3,316 and 469 individuals, which was considered sufficiently large to detect significant associations in our regression analysis.

**Figure 3.**
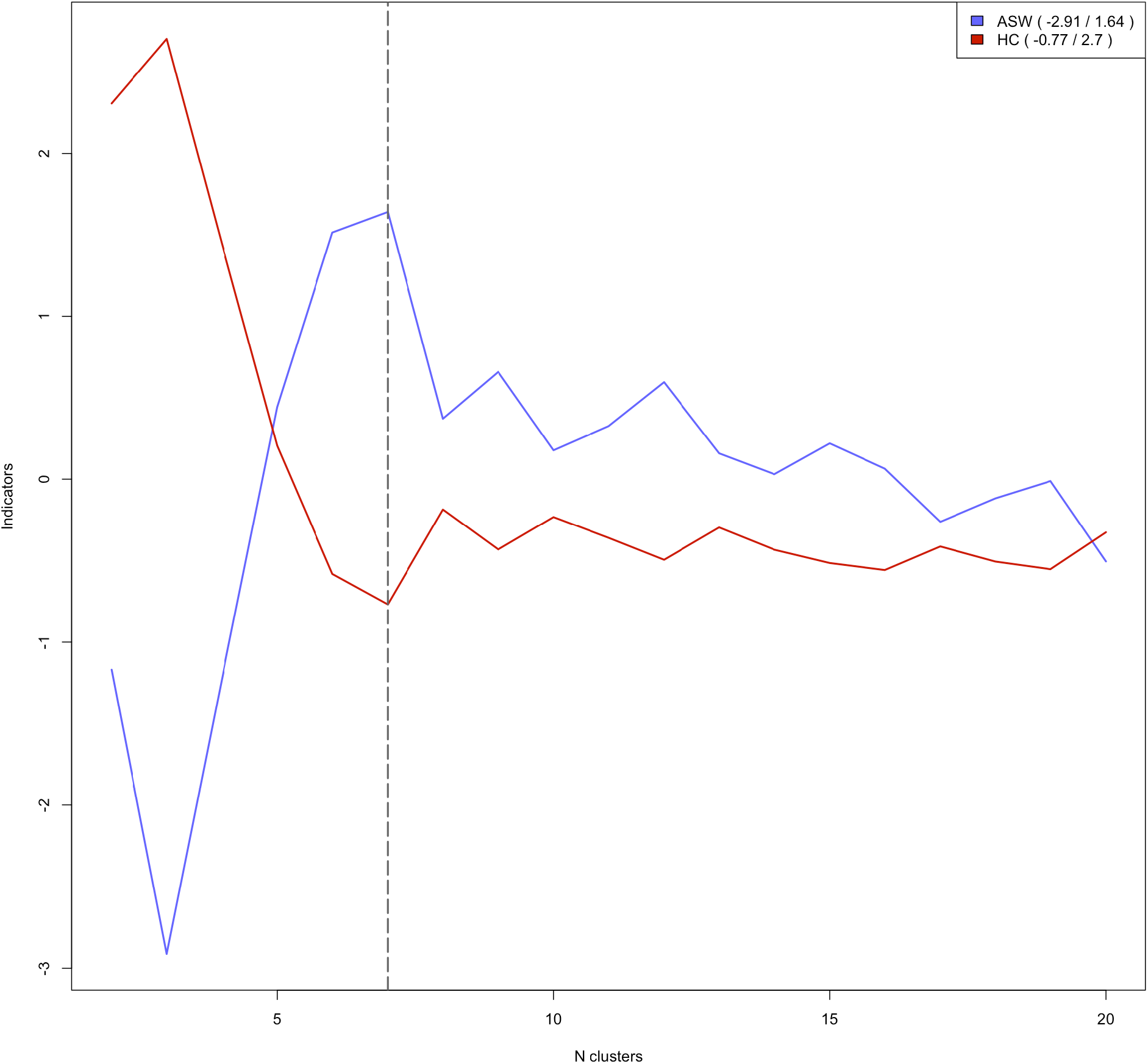
– Standardized cluster quality indicators for a range of cluster solutions. This figure graphically visualizes two cluster quality measures (Average Silhouette Width, ASW and Hubert & Levin C index, HC) for a chosen range of partitions (2-20 clusters) obtained using the Partition Around Medoids (PAM) clustering algorithm to group transition-to-retirement trajectories. Both measures were z-standardize to improve the plot’s readability. When identifying the highest quality cluster solution, the goal is to maximize the ASW and minimize HC. Based on this guideline, we selected 7 clusters as the final partition use to characterize transition-to-retirement trajectories and examine their association with later-life health.

**Figure 4.**
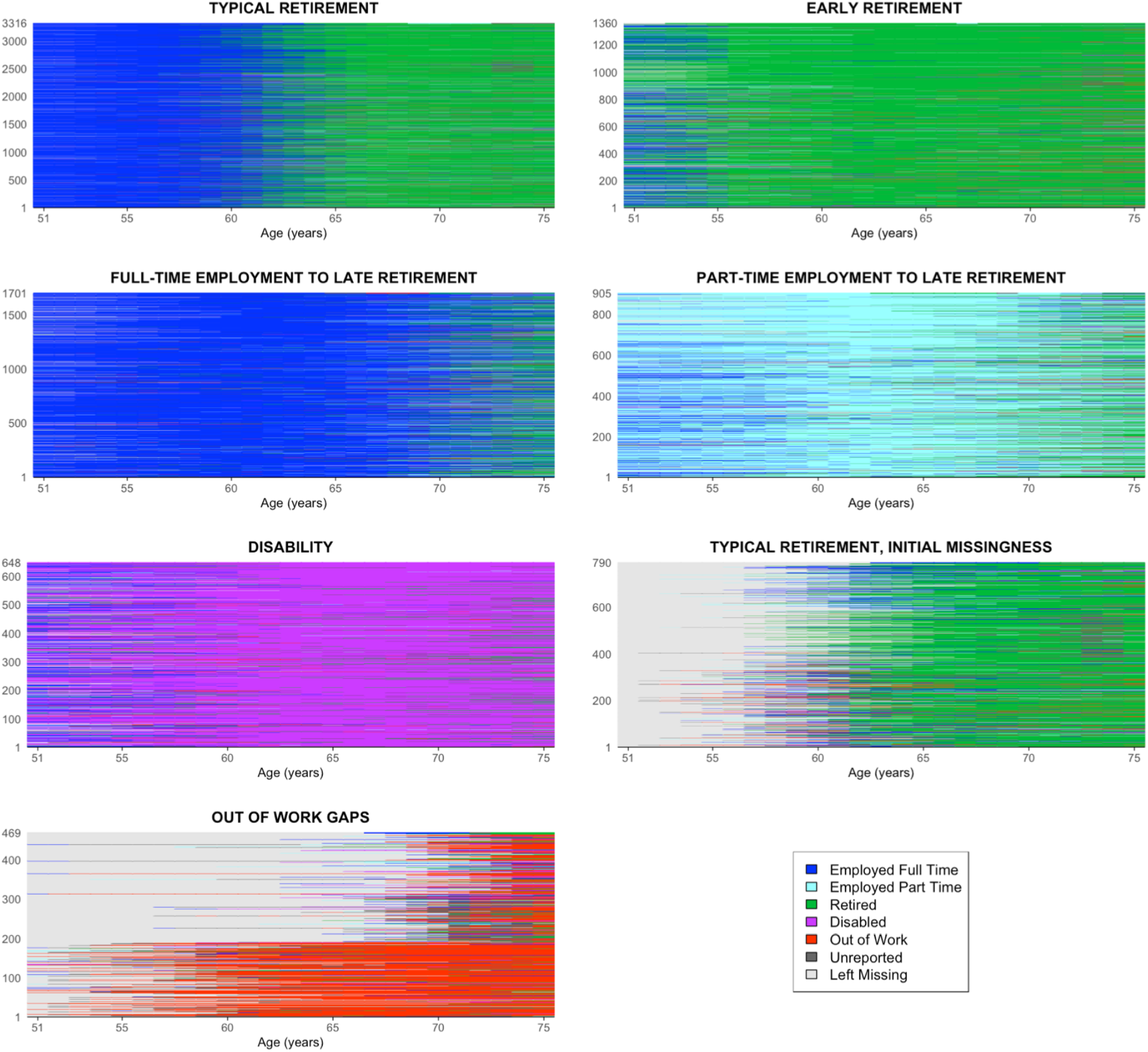
– Sequence Index Plots by cluster. This figure represents sequence index plots for the 7 clusters characterizing transition-to-retirement trajectories among 9,189 Health and Retirement Study participants. Clusters were identified with the goal of grouping similar transition-to-retirement trajectories based on the dissimilarity measure obtained by applying the Hamming distance. Based on this measures, clusters were generated using the Partition Around Medoids (PAM) algorithm. Cluster size ranged between 3,316 participants (“Typical Retirement” cluster) and 469 participants (“Out of work gaps”).

Figure 4 illustrates the sequence index plots for the 7 identified clusters: 1) typical retirement (retirement between age 60-65; N=3,316); 2) early retirement (retirement before age 60; N=1,360) ; 3) full time employment to late retirement (N=1,701); 4) part-time employment to late retirement (N=905); 5) disability (N=648); 6) typical retirement, initial missingness (N=790); 7) out of work gaps (out of work gaps towards the end of the trajectory, with unreported initial employment history; N=469).

The manuscript’s supplement presents two other visualization plots commonly used in sequence analysis to display clusters. The modal state plots (Figure S1) are bar graphs showing the most frequent state at each time point for each cluster. The state distribution plots (also known as chronograms, Figure S2) show the proportions of states at each time point for each cluster. Although these two representations lead to a loss of individual information compared to the sequence index plots, they can be helpful visualization strategies to summarize cluster characteristics^8^.

### Step 4: Regression analysis

We used the identified clusters to evaluate the association between transition-to-retirement trajectories (age 51 - 75) and self-rated health (ages 76 or 77) using linear regression analysis. Self-rated health ranged from 1 to 5, with higher values indicating poorer health. When conducting this kind of analysis, an important consideration regards the choice of covariates.

Confounders must temporally precede the exposure period (25 years between age 51-75) given that adjusting for post-exposure variables may induce bias^36^. We chose to include birth year, gender, race and ethnicity, birth place and school years as potential confounders affecting the relationship between transition to retirement trajectories and health.

Figure 5 shows the association between transition-to-retirement clusters and self-rated health. Compared to the “typical retirement” trajectory (reference), the “early retirement” trajectory was associated with slightly worse self-rated health. The two late retirement trajectories were associated with better physical health, while the disability trajectory was associated with poorest health compared to the reference group. Not reporting employment history before retirement was not associated with a difference in self-rated health, while those in the “out of work gaps” cluster had slightly worse self-rated health than the reference group. We use these results as a didactic example and acknowledge that health experiences before the transition to retirement (which were not included in our confounder set) or during the transition to retirement (which currently sequence analysis approaches cannot incorporate) may be important confounders to consider when interpreting these results.

**Figure 5.**
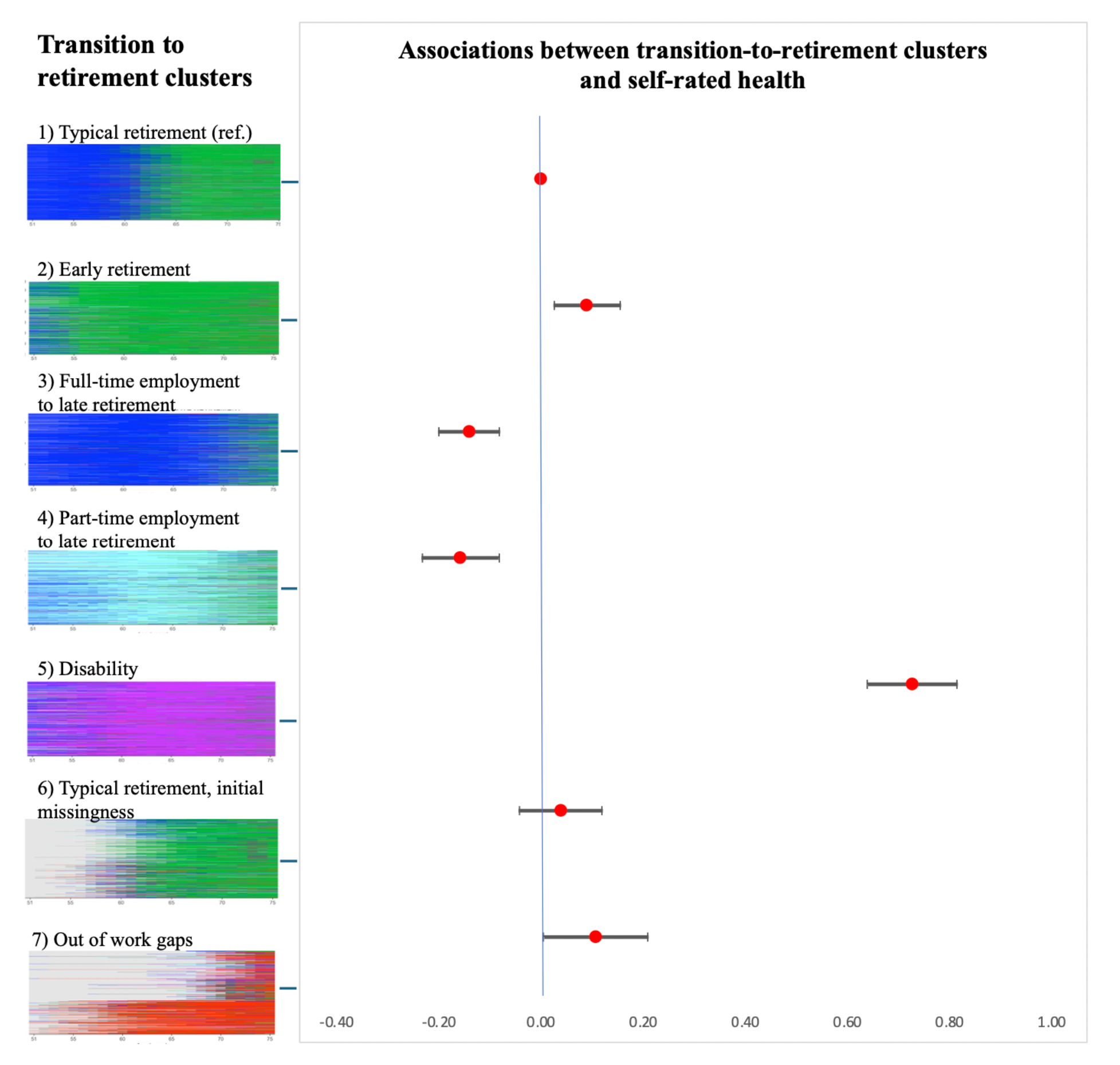
– Association between transition-to-retirement clusters and self-reported health. This figure shows coefficient plots for the association between transition-to-retirement clusters (age 51-75) and self-rated health (age 76 or 77) in the Health and Retirement Study. Self-rated health ranged between 1 and 5, with higher values indicating worse health. Associations were estimated using linear regression, with typical retirement (the most common trajectory) as the reference group. The regression model was adjusted for a set of confounders that preceded the transition-to-retirement trajectories: gender, race and ethnicity, birth place and school years.

### Available software for sequence analysis

Sequence and cluster analysis can be implemented both in Stata and R. The main packages for sequence analysis are SQ and SADI in Stata^37,38^, and TraMineR in R^39^. Although the options provided by the different software packages are similar, we chose to use R for the sequence and cluster analysis in our didactic paper, as its sequence and cluster analysis packages are more complete and recently updated compared to the ones offered in Stata. For example, the Stata packages do not include some of the recently developed dissimilarity measures, and Stata does not allow performing PAM clustering using a dissimilarity matrix. In addition, the R packages have the advantage of being faster, a desirable feature when dealing with very large datasets. In the supplement to the manuscript (Table S5), we illustrate the main available software packages and options for each of the steps of sequence analysis.

## DISCUSSION

Sequence analysis represents an innovative approach for lifecourse epidemiology research, increasingly applied to uncover trajectories of socio-economic determinants of health over long periods of time or for understanding health trajectories^16,40^. In this paper, we introduced sequence analysis for epidemiologic research and guided researchers through the different steps of this method and its application. First, we defined sequences and described the algorithmic approach used to compare pairs of trajectories. Second, we illustrated criteria for choosing a dissimilarity measure, used to compare all sequences in the data. Finally, we introduced clustering with the goal of grouping similar sequences, presented the main clustering algorithms and cluster quality indicators currently used in sequence analysis. Each of these steps was applied using a real-world example, aimed at creating transition-to-retirement trajectories in the HRS and estimating their association with later-life health.

Sequence analysis has several advantages compared to other methods used to analyze longitudinal data. First, it considers trajectories as a whole, and allows for characterization of processes that evolve over time. This is particularly promising in the field of lifecourse epidemiology, which has as its main goal to understand and document how lifecourse processes influence health. Second, as a data reduction technique it facilitates summarizing a very large number of individual trajectories and identifying relevant patterns in timing, order, and duration of events, while maintaining meaningful heterogeneities. Finally, it is a flexible, non-parametric approach, easily tailored to the specific research question of interest^8^.

Despite its potential and advantages, the application of sequence analysis can be challenging, particularly for those implementing this approach for the first time. Each of its steps - from data cleaning to create individual trajectories, to selecting an approach to calculate sequence dissimilarity, to identifying clustering algorithms and the final number of clusters - involves key decisions from myriad existing and emerging options - a time consuming and, at times, confusing process. Indeed, sequence analysis has been criticized for the volume of user set parameters and resulting risk for arbitrary decision-making^8,41^. In this manuscript, we present a principled approach to each of these decisions, drawing from the most recommended. We are aware that sequence analysis is an evolving field, and that more options and extensions of this method will be available, which we will incorporate into future work.

## CONCLUSION

This paper represents a comprehensive guide to the application of modern sequence analysis tools in the field of lifecourse epidemiology. Sequence analysis is a promising tool both for evaluating the relationship between life course factors and health, and for summarizing and describing healthcare trajectories, which can help identify the most vulnerable patients and provide clinical recommendations. We anticipate that sequence analysis will be increasingly used in healthcare research, and we hope that our paper will serve as a useful tool to guide researchers through its implementation.

## Data Availability

All data produced in the present study are available upon reasonable request to the authors.

## ACKNOWLEDGEMENTS

We would like to thank M. Studer, C. Brzinsky Fay, K. Emery and L. Unterlerchner for their methodological and software advice on sequence analysis. We would like to acknowledge the Sequence Analysis Association as a valuable resource and network for the development of this paper.

