## Supplemental Tables and Figures for "Sequence Analysis as an approach to characterize variables that unfold over time: implementation and practical considerations for epidemiologists"

### SUPPLEMENTAL MATERIAL

#### Table S1 – Glossary of terms

| Term | Synonyms | Definition |
|---|---|---|
| sequence | trajectory | Ordered list of elements. In the social science, a sequence represents a ordered succession of lifecourse events over a defined time period (e.g., someone's employment status in each month during one year) |
| state | event; element | Mutually exclusive category experienced by one individual at each time point; the ordered succession of states constitutes a sequence, or trajectory |
| cost | - | When comparing a pair of sequences, a cost is a "penalty" assigned to each transformation operation necessary to transform one sequence into another. In Optimal Matching and its extensions, individual costs for each transformation operation are summed to.obtain total costs corresponding to a quantitative masure of sequence dissimilarity |
| dissimilarity | distance | Measure obtained in sequence analysis, through an algorithmic procedure, to quantitatively compare sequences. Lower dissimilarity values indicate more similar sequences, while higher dissimilarity values indicate less similar sequences. |
| Optimal Matching | - | Most commonly used algorithm to compare trajectories in sequence analysis. It uses an iterative minimization procedure to find the distance between every pair of sequences in a sample. |
| cluster | group; pattern | Set of similar sequences, grouped together based on their dissimilarity measure. A "good quality" cluster should be as homogenous as possible within itself, as as distinct as possible from the other clusters. |
| cluster quality measure | cluster quality indicator | Statistic used to compare cluster quality across different partitions, each composed of a different number of clusters. |

This table defines some of the main terms specific to sequence analysis, and often used in our manuscript. Because some of the terms are using interchangeably (e.g., sequence and trajectory; distance and dissimilarity), this table lists synonyms for each of the relevant term in column # 2, with the goal of improving the manuscript's clarity and readability.

**Table S2 - Substitution cost matrix based on transition frequencies between states**

| States | Employed Full-Time | Employed Part-Time | Retired | Disabled | Out of Work | Unreported | Left Missing |
|---|---|---|---|---|---|---|---|
| **Employed Full-Time** | 0.00 | 1.96 | 1.95 | 1.99 | 1.98 | 1.91 | 1.96 |
| **Employed Part-Time** | 1.96 | 0.00 | 1.95 | 1.99 | 1.99 | 1.90 | 1.98 |
| **Retired** | 1.95 | 1.95 | 0.00 | 1.97 | 1.97 | 1.67 | 1.97 |
| **Disabled** | 1.99 | 1.99 | 1.97 | 0.00 | 1.99 | 1.95 | 1.99 |
| **Out of Work** | 1.98 | 1.99 | 1.97 | 1.99 | 0.00 | 1.72 | 1.99 |
| **Unreported** | 1.91 | 1.90 | 1.67 | 1.95 | 1.72 | 0.00 | 1.98 |
| **Left Missing** | 1.96 | 1.98 | 1.97 | 1.99 | 1.99 | 1.98 | 0.00 |

This table displays the symmetric, square substitution cost matrix obtained by setting costs as inversely proportional to the transitions between states in our applied example dataset, representing transition-to-retirement trajectories (from age 51 to age 75) in the Health and Retirement Study (HRS). The “states” used to categorize employment status at each time point are displayed on the top row and left column. Substitution costs range between 0 (substituting a state with itself) and 2 (substituting two states when no transition between these two is observed). Within this range, lower costs (e.g., cost of substituting “out of work” with “unreported” , and vice-versa) correspond to more frequent transitions, while higher costs (e.g., cost of substituting “disabled” with “employed full-time”, and vice-versa) correspond to less frequent transitions.

**Table S4 – Dissimilarity matrix**

| | 1 | 2 | 3 | 4 | 5 | 6 | 7 | 8 | 9 | 10 | 11 | 12 | 13 | 14 | 15 |
|---|---|---|---|---|---|---|---|---|---|---|---|---|---|---|---|
| **1** | **0.00** | 49.07 | 15.78 | 31.18 | 29.20 | 49.06 | 32.80 | 32.31 | 28.84 | 47.18 | 34.77 | 49.28 | 49.18 | 43.69 | 31.13 |
| **2** | 49.07 | **0.00** | 43.35 | 33.53 | 47.44 | 43.69 | 49.37 | 38.99 | 43.38 | 31.80 | 41.57 | 21.87 | 21.77 | 37.39 | 47.41 |
| **3** | 15.78 | 43.35 | **0.00** | 27.11 | 44.97 | 43.16 | 48.58 | 24.37 | 20.84 | 41.49 | 48.60 | 49.36 | 49.31 | 43.84 | 44.95 |
| **4** | 31.18 | 33.53 | 27.11 | **0.00** | 43.28 | 43.28 | 46.88 | 30.21 | 32.93 | 41.65 | 46.90 | 27.55 | 27.23 | 32.19 | 45.11 |
| **5** | 29.20 | 47.44 | 44.97 | 43.28 | **0.00** | 49.35 | 25.36 | 44.36 | 43.03 | 49.31 | 13.62 | 49.64 | 49.55 | 48.92 | 33.50 |
| **6** | 49.06 | 43.69 | 43.16 | 43.28 | 49.35 | **0.00** | 49.05 | 38.86 | 28.81 | 35.84 | 49.12 | 49.71 | 49.67 | 46.25 | 33.21 |
| **7** | 32.80 | 49.37 | 48.58 | 46.88 | 25.36 | 49.05 | **0.00** | 46.40 | 46.63 | 49.09 | 31.19 | 49.61 | 49.52 | 47.07 | 27.38 |
| **8** | 32.31 | 38.99 | 24.37 | 30.21 | 44.36 | 38.86 | 46.40 | **0.00** | 28.44 | 41.15 | 46.00 | 49.10 | 48.79 | 38.21 | 47.22 |
| **9** | 28.84 | 43.38 | 20.84 | 32.93 | 43.03 | 28.81 | 46.63 | 28.44 | **0.00** | 41.52 | 46.67 | 49.41 | 49.34 | 43.42 | 35.14 |
| **10** | 47.18 | 31.80 | 41.49 | 41.65 | 49.31 | 35.84 | 49.09 | 41.15 | 41.52 | **0.00** | 49.38 | 37.81 | 37.76 | 45.23 | 47.36 |
| **11** | 34.77 | 41.57 | 48.60 | 46.90 | 13.62 | 49.12 | 31.19 | 46.00 | 46.67 | 49.38 | **0.00** | 49.65 | 49.56 | 48.71 | 37.19 |
| **12** | 49.28 | 21.87 | 49.36 | 27.55 | 49.64 | 49.71 | 49.61 | 49.10 | 49.41 | 37.81 | 49.65 | **0.00** | 3.94 | 21.58 | 45.61 |
| **13** | 49.18 | 21.77 | 49.31 | 27.23 | 49.55 | 49.67 | 49.52 | 48.79 | 49.34 | 37.76 | 49.56 | 3.94 | **0.00** | 17.64 | 45.52 |
| **14** | 43.69 | 37.39 | 43.84 | 32.19 | 48.92 | 46.25 | 47.07 | 38.21 | 43.42 | 45.23 | 48.71 | 21.58 | 17.64 | **0.00** | 40.29 |
| **15** | 31.13 | 47.41 | 44.95 | 45.11 | 33.50 | 33.21 | 27.38 | 47.22 | 35.14 | 47.36 | 37.19 | 45.61 | 45.52 | 40.29 | **0.00** |

This table displays the top-left corner of the dissimilarity (or distance) matrix obtained by applying sequence analysis to transition-to-retirement trajectories in the Health and Retirement study. After setting substitution costs between states (Table S1), we applied the Hamming distance. This method calculates sequence dissimilarity for each pair of sequences by summing the substitution costs of replacing states that differ between pairs of sequences at each time point across the whole chosen period. Dissimilarity values between all pairs of sequences in the dataset are represented by the dissimilarity (or distance) matrix, a symmetric, square matrix where the number of rows and columns correspond to the number of individuals in the data (in our case, a 9,189*9,189 matrix). In this table, we show dissimilarity measures for the first 15 participants in the dataset. Higher values correspond to more dissimilar sequences (e.g., participants 1 and 2), while lower values correspond to more similar sequences (e.g., participants 12 and 13).

### Table S4 – Cluster quality measures for hierarchical and PAM clustering

**Hierarchical Clustering**

| | **partition #1** | | **partition #2** | | **partition #3** | | **partition #4** | |
|---|---|---|---|---|---|---|---|---|
| | N groups | stat | N groups | stat | N groups | stat | N groups | stat |
| ASW | 10 | 0.324 | 9 | 0.320 | 5 | 0.317 | 8 | 0.314 |
| CH | 3 | 1410 | 6 | 1344 | 4 | 1338 | 5 | 1268 |
| HC | 13 | 0.102 | 12 | 0.107 | 10 | 0.108 | 9 | 0.111 |

**Partition Around Medoids (PAM)**

| | **partition #1** | | **partition #2** | | **partition #3** | | **partition # 5** | |
|---|---|---|---|---|---|---|---|---|
| | N groups | stat | N groups | stat | N groups | stat | N groups | stat |
| ASW | 7 | 0.358 | 6 | 0.354 | 9 | 0.323 | 12 | 0.321 |
| CH | 6 | 1525 | 5 | 1441 | 7 | 1395 | 2 | 1365 |
| HC | 7 | 0.095 | 6 | 0.106 | 16 | 0.107 | 19 | 0.108 |

This table displays three of the most common cluster quality indicators (Average Silhouette Width, ASW; Calinski–Harabasz index, CH; Hubert & Levin C index, HC) for two clustering algorithms (hierarchical clustering and partition around medoids), applied to grouping transition-to-retirement trajectories in the Health and Retirement Study based on sequence dissimilarity. For each of the clustering algorithms, we display cluster quality for the four best partitions. Comparing cluster quality indicators between different clustering algorithm can help select which algorithm will be used in the final analysis. Higher ASW and CH and lower HC indicate better cluster quality. Because PAM clustering produces, overall, better quality partitions, we decided to use this algorithm in our didactic example characterizing transition-to-retirement trajectories. Mathematical details on each of the measures can be found in the Weighted Cluster manual.

**Figure S1 – Sequence modal plots by cluster**

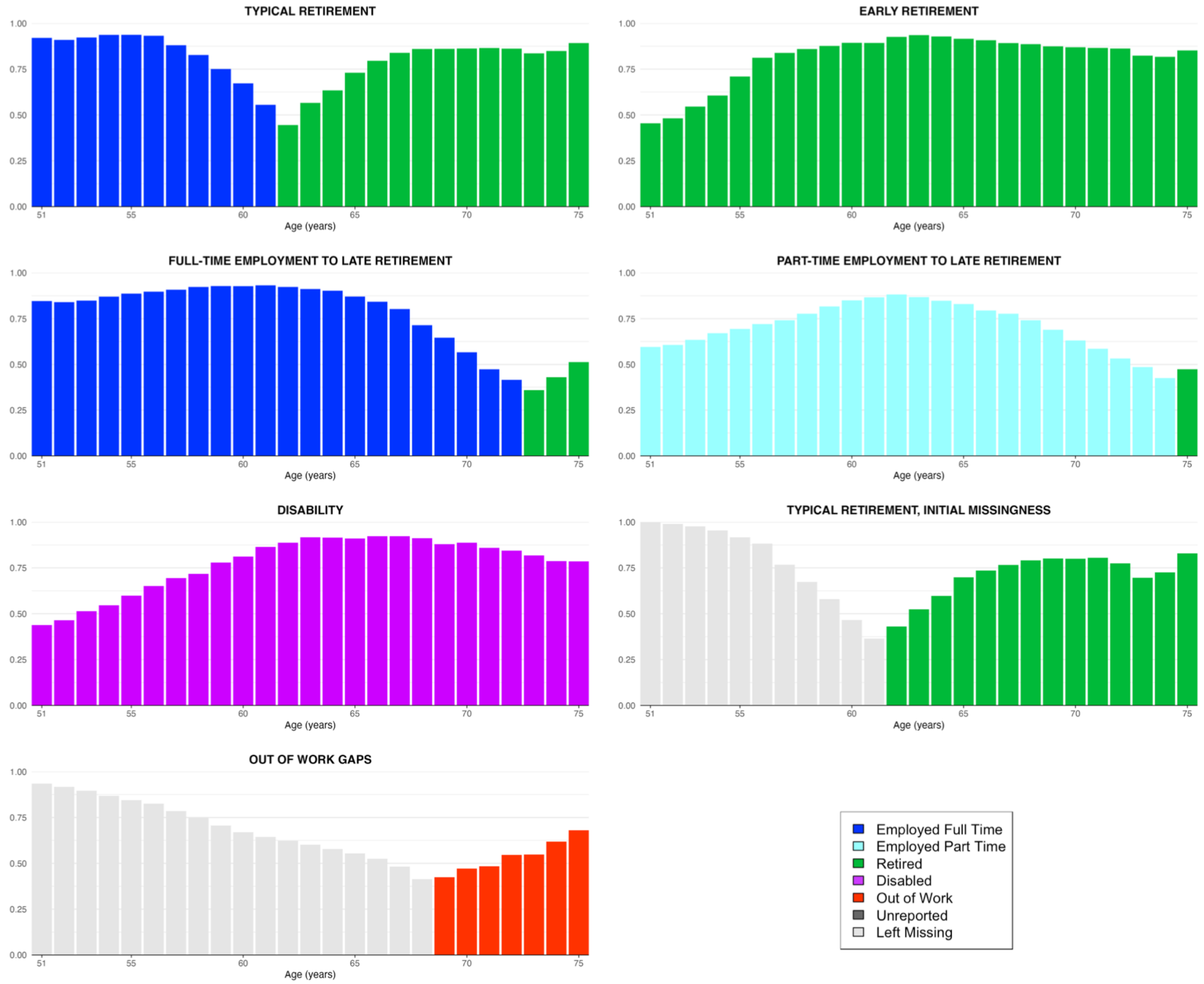


This figure displays sequence modal plots for the 7 clusters obtained in our applied example, which compares and groups transition-to-retirement trajectories of 9,189 Health and Retirement Study participants. The sequence modal plot shows the most common state (mode) at each time point (in our example, each year between age 51 and age 75, x-axis). The y-axis shows the relative frequencies of the modal state at each time point. Although sequence modal plots lead to a loss of information compared to sequence index plots (Figure 4 of the manuscript), they are useful summary graphics commonly used in sequence analysis.

### Figure S2 – State distribution plots (chronograms) by cluster

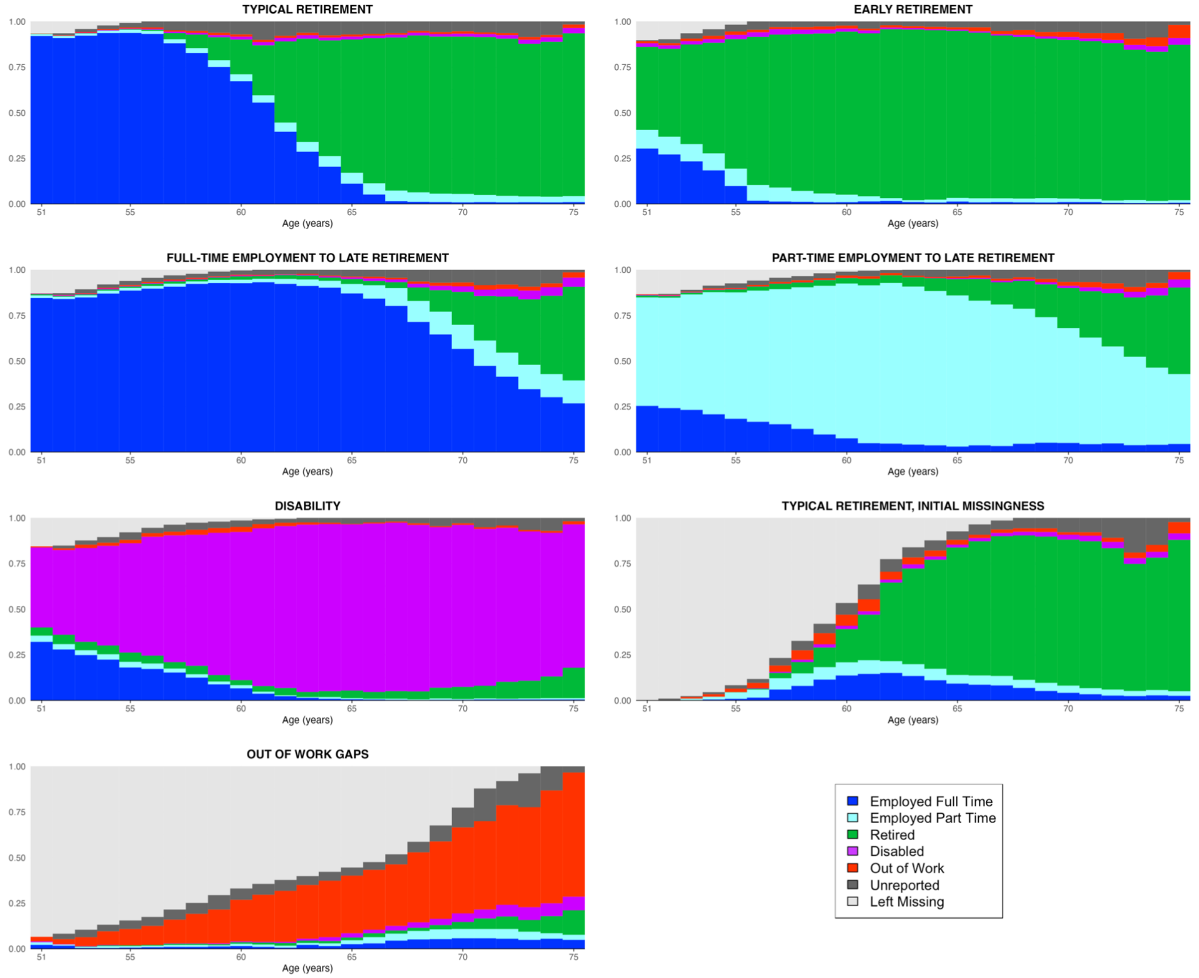


This figure displays sequence distribution plots (also known as chronograms) for the 7 clusters obtained in our applied example, which compares and groups transition-to-retirement trajectories of 9,189 Health and Retirement Study participants. State distribution plots show the proportion (x-axis) of states at each time point (age from 51-75 in our example, y-axis). In the “typical retirement” cluster, for example, about 90% of participants are employed full time in the first few years of the trajectory. This proportion gradually decreases after age 55, while the proportion of retired participants increases. State distribution plots are more detailed graphical representations than modal plots, but still lead to a loss of information compared to sequence index plots (Figure 4 of the manuscript).

**Figure S3 – Transition-to-retirement index plots by clusters obtained using OM algorithm**

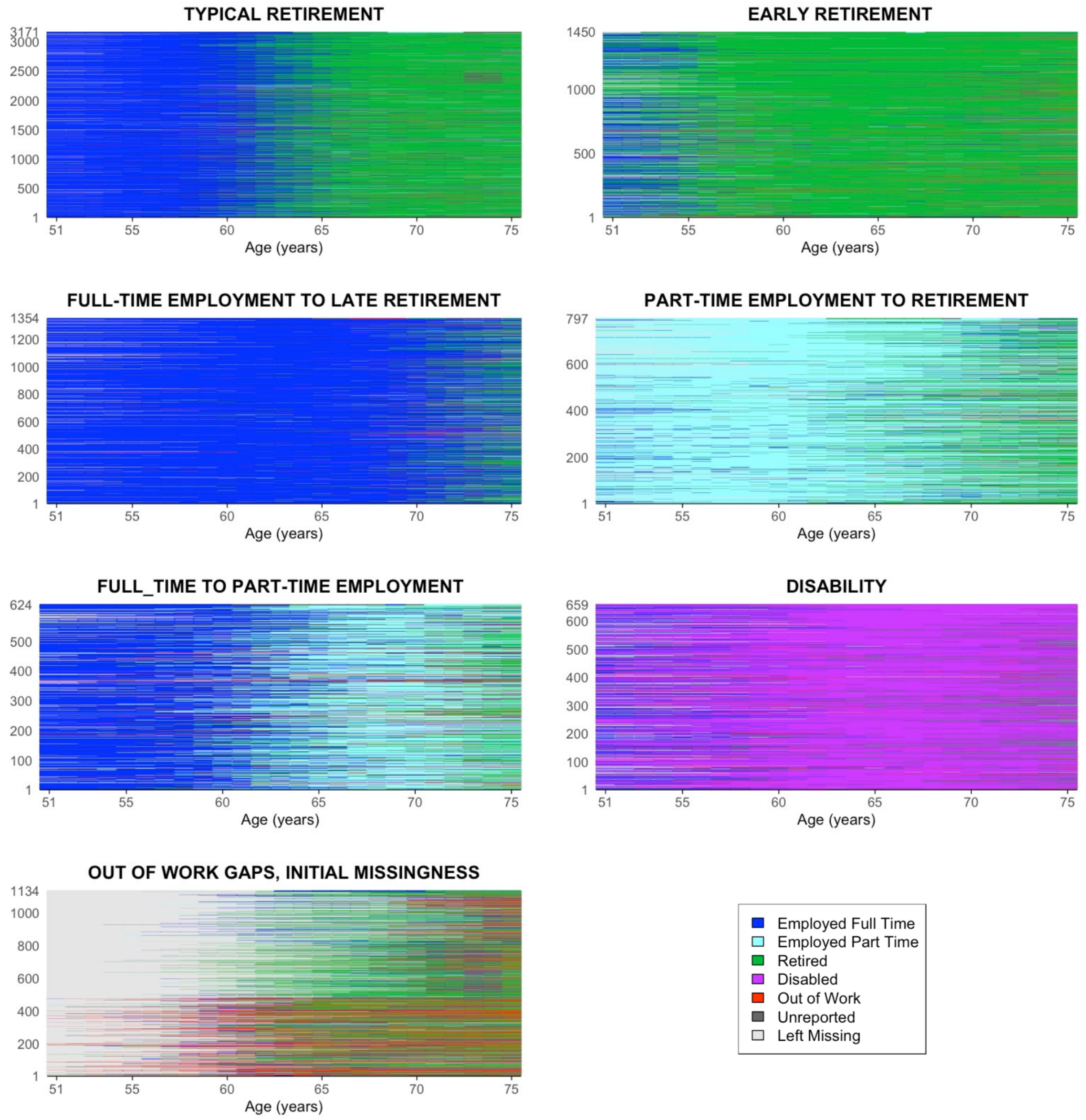


This figure represents sequence index plots for clusters obtained in a sensitivity analysis applying the Optimal Matching algorithm to comparing and grouping transition-to-retirement trajectories in the Health and Retirement Study. While the Hamming distance, used in our didactic example to calculate sequence dissimilarity, prioritizes the timing of states, the Optimal Matching algorithm tends to prioritize duration of states.

**Table S5 – Software packages and functions for sequence analysis**

| Steps of sequence analysis | Task | R | STATA |
|---|---|---|---|
| **1) Creating and visualizing individual sequences** | Sequence definition | package: TraMineR, function: seqdef | package: SQ, function: sqset |
| | Options for treating left, internal and right missing data | package: TraMineR, function: seqdef; options: left (), gaps(), right() | Left missing values and gaps need to be coded as separate states. Right missing values are deleted if a variable sequence length is specified when generating the dissimilarity matrix. |
| | Imputation of sequence data (Halpin, 2012) | package: seqimpute | package: MICT |
| | Sequence Index plot | package: TraMineR, function: seqIplot | package: SQ, function: sqindexplot |
| | Sequence modal plot | package: TraMineR, function: seqmsplot | package: SQ, function: sqmodalplot |
| | Chronogram (state distribution plot) | package: TraMineR, function: seqdplot | package: SADI, command: sdchronogram |
| **2) Quantifying trajectory dissimilarity** | Choose costs | package: TramineR, function: seqdist, options: | package: SADI, function: [depends on the dissimilarity matrix], option: subsmat(substitution matrix name) |
| | Choose dissimilarity measure | package: TramineR, function: seqdist, options: HAM (Hamming); OM (Optimal Matching) | package: SADI, function: oma (Optimal Matching), sdhamming (Hamming) |
| **3) Performing cluster analysis to group similar sequences** | Partition Around Medoids (PAM) clustering | package: WeightedCluster, function: wcKMedoids | No available option |
| | Hierarchical, agglomerative clustering | function: hclust | package: clustermat |
| | Cluster quality indicators | package: WeightedCluster, functions: wcKMedoids (for PAM); as.clustrange (for hierarchical clustering) | packages: dudahart (Duda-Hart stopping rule); silhouette (Average Silhouette Width); calinski (Calinski-Harabasz pseudo-F for stopping rules) |

This table shows the main R and STATA packages, functions and options for each of the tasks included in three steps of sequence analysis. Despite many dissimilarity measures being available, we only display options for Hamming and Optimal Matching, which are used in our applied example. Similarly, for the clustering steps, we show the most commonly used algorithms (hierarchical clustering and PAM). More options are described in the TramineR, SADI and WeightedCluster packages.
